# Short-term Neuropsychiatric Outcomes and Quality of Life in COVID-19 Survivors

**DOI:** 10.1101/2020.09.23.20190090

**Authors:** Raúl Méndez, Vicent Balanzá-Martínez, Sussy C. Luperdi, Itziar Estrada, Ana Latorre, Paula González-Jiménez, Laura Feced, Leyre Bouzas, Katheryn Yépez, Ana Ferrando, David Hervás, Enrique Zaldívar, Soledad Reyes, Michael Berk, Rosario Menéndez

**Author notes:** Equal contribution (Raúl Méndez, Vicent Balanzá-Martínez, Sussy C. Luperdi). **Corresponding author:** Raúl Méndez Servicio de Neumología. Hospital Universitario y Politécnico La Fe, Avda. Fernando Abril Martorell 106, 46026 Valencia. Spain.

## Abstract

**Background:** The general medical impacts of coronavirus (COVID-19) are increasingly appreciated. However, its impact on neurocognitive, psychiatric health and quality of life (QoL) in survivors after the acute phase is poorly understood. We aimed to evaluate neurocognitive function, psychiatric symptoms, and QoL in COVID-19 survivors shortly after hospital discharge.

**Methods:** This was a cross-sectional analysis of a prospective study of hospitalized COVID-19 survivors followed-up for 2 months after discharge. A battery of standardized instruments evaluating neurocognitive function, psychiatric morbidity, and QoL (mental and physical components) was administered by telephone.

**Findings:** Of the 229 screened patients, 179 were included in the final analysis. Among survivors, the prevalence of moderately impaired immediate verbal memory and learning was 38%, delayed verbal memory (11.8%), verbal fluency (34.6%), and working memory (executive function) (6.1%), respectively. Moreover, 58.7% of patients had neurocognitive impairment in at least one function. Rates of positive screening for anxiety were 29.6%, depression (26.8%), and post-traumatic stress disorder (25.1%) respectively. In addition, 39.1% of the patients had psychiatric morbidity. Low QoL for physical and mental components was detected in 44.1% and 39.1% of patients, respectively. Delirium and stress-related symptoms increased approximately 4-fold the odds of developing neurocognitive impairment. Female gender and neurocognitive impairment diagnosis were related with an increase of 2.5 and 4.56- fold odds respectively of psychiatric morbidity.

**Interpretation:** Hospitalized COVID-19 survivors showed a high prevalence of neurocognitive impairment, psychiatric morbidity, and poor QoL in the short-term. It is uncertain if these impacts persist over the long-term.

## INTRODUCTION

The severe acute respiratory syndrome coronavirus 2 (SARS-CoV-2) was identified in December 2019 (1). As of September 22, SARS-CoV-2 caused more than 31 million cases of coronavirus disease and over 965.000 deaths worldwide (2). Efforts by the international community are focusing on decreasing the virus transmission as well as reducing its mortality. Despite the considerable mortality rate, the vast majority of patients survive COVID-19, however potential systemic sequelae are poorly understood and may be an additional global public health issue (3).

Pulmonary fibrosis, cardiovascular damage, and neurological impairment are among the potential sequelae of COVID-19 in survivors. It is known that many human coronaviruses display neurotropism, such as HCoV-OC43, SARS-CoV-1, and the Middle East respiratory syndrome CoV (MERS-CoV) (4). Psychiatric and neurological presentations have been found in the acute and post-illness stage in other severe coronavirus infections (5). Nevertheless, most of the studies were of either low or medium quality and focused on the acute phase of infection (5).

During hospitalization more than a half of COVID-19 patients have neurological symptoms, and their persistence after the acute phase is currently under study (6-8). The most frequently observed neurological symptoms are headache, dizziness, fatigue, anorexia, ageusia, and anosmia (9). Of note, neurocognitive deficits in the post-acute stage of COVID-19 and other coronavirus infections are insufficiently described (10). Only preliminary data support the neurological and neuropsychiatric impact in COVID-19 patients. In a national surveillance study with voluntary case notification, Varatharaj et al reported that 31% of patients had an altered mental status including neuropsychiatric and neurocognitive symptoms in the acute phase (11). To the authors’ knowledge, there are no previous studies in COVID-19 simultaneously evaluating neurocognitive function, psychiatric symptoms (such as stress-related symptoms, including depression, anxiety, and post-traumatic stress disorder [PTSD]), and quality of life (QoL) following the acute phase in survivors.

We hypothesized that COVID-19 survivors may present with persistent impaired cognition, psychiatric symptoms, and poorer QoL. Severity at admission, hypoxemia, systemic inflammation, and respiratory support during hospitalization may be associated with neuropsychiatric and QoL impairment in COVID-19. We thus conducted a cross-sectional cohort study in COVID-19 survivors post hospitalization to assess the neuropsychiatric and QoL consequences 2 months after hospital discharge.

## METHODS

### Design and Participants

We conducted a cross-sectional analysis of a prospective cohort study in a large tertiary care hospital in Valencia, Spain. All hospitalized patients with COVID-19 were diagnosed with reverse transcription polymerase chain reaction (RT-PCR) of nasopharyngeal swab or sputum samples. Exclusion criteria included patients aged ≥85 or <18 years, non-Spanish speaking subjects, nursing-home residents, pre-existing dementia, pre-existing or cognitive decline under evaluation, previous brain injury with cognitive sequelae, current alcohol/substance use disorder (except for nicotine), and previously diagnosed major psychiatric disorders. All screened patients required hospitalization between March 8 and April 25 2020 in the Department of Pneumology and/or the Intensive Care Unit (ICU). Survivors were referred to the COVID-19 outpatient clinic in the Pneumology Department for clinical control where they were invited to participate in the study. All patients provided written informed consent to participate in the study. The Biomedical Research Ethics Committee of La Fe University and Polytechnic Hospital reviewed and approved the study (2020-280-1). The hospital discharge dates were between March 23 to July 7 and outpatient clinic visits were from May 18 to July 24. All recruited patients were contacted by telephone 2 (±1) months from the date of hospital discharge.

### Neuropsychiatric and Quality of Life Assessment

A battery of standardized instruments evaluating neurocognitive function, psychiatric morbidity, and QoL was administered by telephone (eTable 1). The neuropsychiatric evaluation included immediate verbal memory and learning, delayed verbal memory, verbal fluency, and working memory (executive function) for the neurocognitive domain; anxiety, depression, and PTSD were evaluated for the psychiatric domain (stress-related symptoms). The telephone battery was designed by two psychiatrists experienced in neurocognition (12). An independent researcher, blind to clinical data and the study hypothesis, conducted pilot interviews under monitoring of her supervisors before enrolment. The duration of the interview was approximately 20 minutes. Three telephone contact attempts were made with the patients at different times before desisting. The data obtained from the interview were attached to the database with clinical data collected by an independent researcher.

Neurocognitive assessment tests included: i) the Verbal Learning-Immediate and ii) delayed memory subtests from the Subtest Screen for Cognitive Impairment in Psychiatry (SCIP), iii) Animal Naming Test (ANT) from the Controlled Oral Word Association Test (COWAT) for semantic verbal fluency, iv) and the subtest Digit Span backward from the Wechsler Adult Intelligence Scale, Third Edition (WAIS-III) for working memory (executive function). Psychiatric morbidity was assessed with screening questionnaires, including: i) Generalized Anxiety Disorder 7 (GAD-7) for anxiety, ii) Patient Health Questionnaire 2 (PHQ-2) for depression, iii) and 17-item Davidson Trauma Scale (DTS) for PTSD. Finally, for QoL assessment, the Short Form Health Survey 12 item (SF-12, First version) was administered. This QoL test provides two independent scores, a physical component summary (PCS) and a mental component summary score (MCS). Additional information on neuropsychiatric and QoL battery is detailed in the Supplemental File.

### Clinical Data

Relevant variables such as demographics, comorbidities, chronic treatments, laboratory parameters, and radiographic data were recorded. Initial severity was measured with Pneumonia Severity Index (PSI), a classical prognostic score for community-acquired pneumonia, given the absence of COVID-19 specific prognostic scores when the study was designed. Blood oxygen saturation/fraction of inspired oxygen (SpO2/FiO2) at admission, respiratory support (need for supplementary oxygen, high-flow nasal cannula oxygen therapy, continuous positive airway pressure [CPAP], non-invasive mechanical ventilation, and/or invasive mechanical ventilation), and laboratory data during hospitalization were recorded. Clinical outcomes included length of hospital stay, ICU admission, venous thromboembolic event, or delirium, among others. Delirium, defined as an acute, reversible state of confusion, was recorded.

During follow-up visits, a checklist of enduring symptoms was administered. All the symptoms were dichotomized as present or absent. This checklist included: fatigue, dyspnoea (if grade ≥2 on the MRC scale), cough, sputum production, chest pain, dysgeusia/ageusia, anosmia, headache, arthralgia, myalgia, paraesthesia, hypoesthesia, tremors, and memory complaints. Blood extraction for routine analysis and future studies was obtained. Vital signs and anthropometric data were also recorded.

### Outcomes

Neurocognitive domain impairment was pre-defined as moderate/severe impairment of any of the four neuropsychological tests. In the same way, psychiatric morbidity was pre-defined as positive screening in any of the three questionnaires assessing stress-related symptoms. This definition was used in previous studies of acute lung injury and neurocognitive evaluation (13). The reference values and cut-off points for the QoL questionnaire and for each variable in the neurocognitive and psychiatric domains are detailed in the Supplemental File.

### Statistical Analysis

Data were summarized as N (%) or median [1^st^ quartile, 3^rd^ quartile], as appropriate. Correlations were estimated using Spearman’s rank-correlation. An exploratory analysis with pre-defined risk factors included known risk factors in ARDS such as delirium, use of corticosteroids, mechanical ventilation, length of ICU stay, metabolic alterations (renal and hepatic) along with those previously described in the hypothesis (14, 15). Multivariable Bayesian logistic regressions with weakly regularizing priors, N(0,1), were used to evaluate the association of risk factors with neurocognitive impairment and psychiatric morbidity in COVID-19 survivors. In the neurocognitive impairment model, age and level of education were not included since tests were adjusted for that variables. Association between stress-related symptoms and neurocognitive impairment was assessed with Chi^2^ test. Statistical significance was considered if P<0.05. Statistical analyses were performed using R (version 4.0.2).

## RESULTS

### Recruitment and Patient Baseline Characteristics

A total of 229 patients discharged after COVID-19 hospitalization were followed in the outpatient clinic of the Pneumology Department. Of those, 197 were contacted by telephone, and 179 completed the test battery (Figure 1). The final data analysis was performed in this cohort.

**Figure 1.**
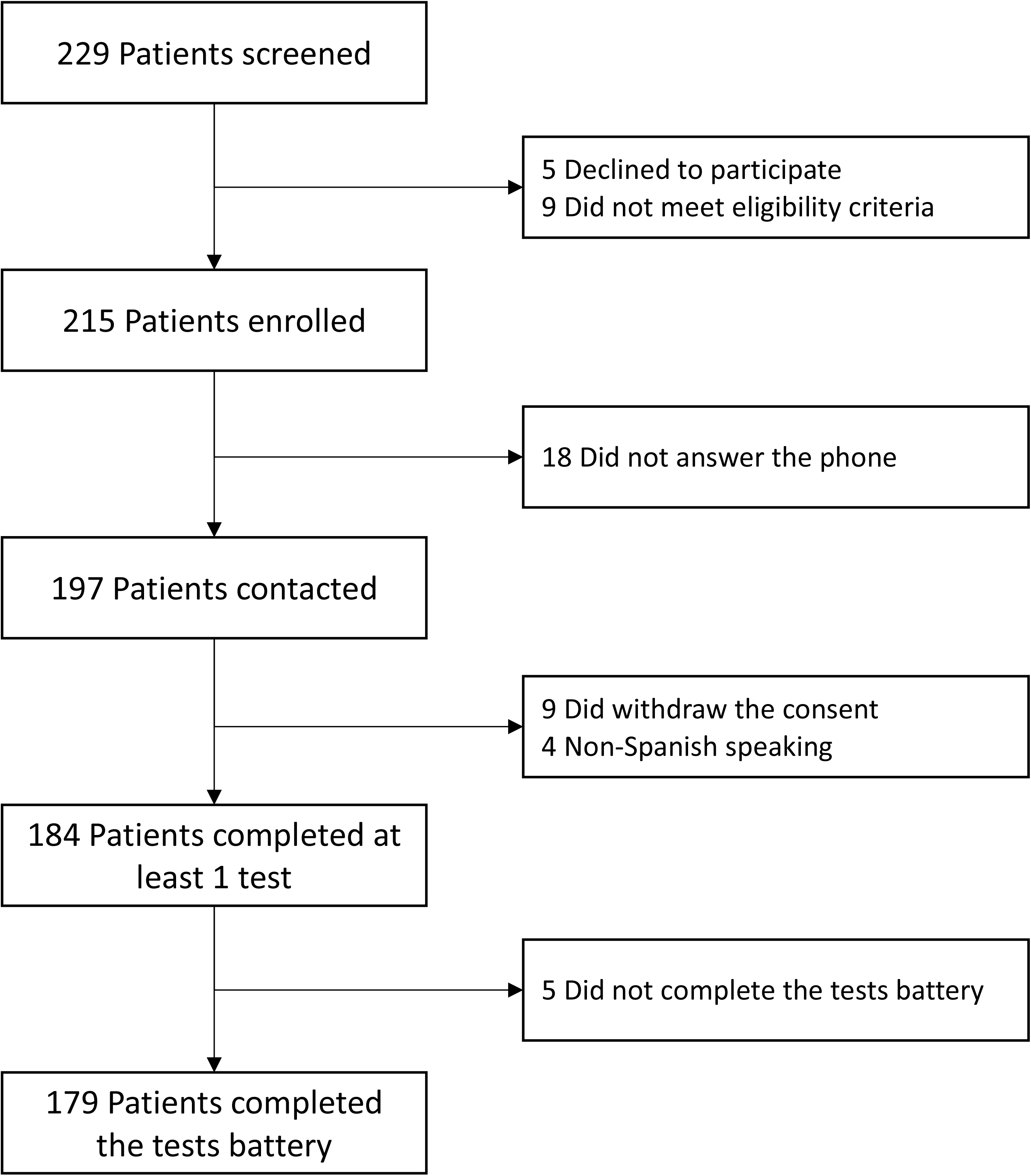
Flowchart. The 179 patients were included in the final analysis.

Table 1 describes the main baseline characteristics of the study sample. Participants’ age ranged from 22 to 81 years and 74 (41.3%) were female. Of the 179 survivors studied, 99 (55.3%) had at least one comorbidity. Only 1.1% of the patients did not complete any formal education and 31.3%, 35.2%, 24.6%, and 7.8% of the patients completed primary, secondary, university, and postgraduate education, respectively. The patients had a median of 7 [6, 10] days from the onset of symptoms to when they were hospitalized. Initial severity measured by PSI score, analytical parameters, respiratory support, treatments, and clinical outcomes during hospitalization also appear on Table 1. No patient had any cerebrovascular accident during the study period.

**Table 1.** Characteristics of the patients at baseline, severity, treatment received, analytical parameters, level of respiratory support, and outcomes.

|  | **Total**<br>**(N=179)** |
| --- | --- |
| Demographics |  |
| Age, yr, median [1<sup>st</sup>, 3<sup>rd</sup> quartile] | 57 [49, 67] |
| Distribution, no. (%) |  |
| <50, yr | 52 (29.1) |
| 50 to 69, yr | 94 (52.5) |
| ≥70, yr | 33 (18.4) |
| Male sex, no. (%) | 105 (58.7) |
| Level of education, yr, median [1<sup>st</sup>, 3<sup>rd</sup> quartile] | 11 [8, 16] |
| Smoking, no. (%) |  |
| Never | 125 (69.8) |
| Former | 44 (24.6) |
| Current | 10 (5.6) |
| Coexisting conditions, no. (%) |  |
| Any | 99 (55.3) |
| Hypertension | 58 (32.4) |
| Diabetes | 29 (16.2) |
| Dyslipidemia | 52 (29.1) |
| Chronic heart disease | 10 (5.6) |
| Chronic renal disease* | 3 (1.7) |
| Chronic liver disease | 3 (1.7) |
| Cancer | 3 (1.7) |
| Chronic respiratory disease | 21 (11.7) |
| Previous medication use, no. (%) |  |
| Antiplatelets | 6 (3.4) |
| Statins | 37 (20.7) |
| ACE inhibitor | 14 (7.8) |
| Angiotensin II-receptor antagonist | 28 (15.6) |
| SpO2/FiO2 at admission, median [1<sup>st</sup>, 3<sup>rd</sup> quartile] | 452.4 [442.9, 461.9] |
| Radiological data at admission |  |
| Lung infiltrates, no. (%) | 177 (98.9) |
| Bilateral infiltrates, no. (%) | 120 (67) |
| Severity |  |
| PSI score, median [1<sup>st</sup>, 3<sup>rd</sup> quartile] | 57 [45, 71] |
| Distribution, no. (%) |  |
| I-III | 169 (94.4) |
| IV-V | 10 (5.6) |
| Analytical parameters |  |
| Peak LDH, UI/L, median [1<sup>st</sup>, 3<sup>rd</sup> quartile] | 317 [258, 436] |
| Peak C-reactive protein, mg/L, median [1<sup>st</sup>, 3<sup>rd</sup> quartile] | 97.2 [45.3, 174.3] |
| Nadir lymphocyte count, cells/mL, median [1<sup>st</sup>, 3<sup>rd</sup> quartile] | 920 [640, 1260] |
| Peak D-dimer, ng/mL, median [1<sup>st</sup>, 3<sup>rd</sup> quartile] | 962 [498, 2102] |
| Treatment, no. (%) |  |
| Hydroxychloroquine, no. (%) | 168 (93.9) |
| Azithromycin, no. (%) | 166 (92.7) |
| Lopinavir/Ritonavir, no. (%) | 76 (42.5) |
| Interferon $\beta$, no. (%) | 26 (14.5) |
| Tocilizumab, no. (%) | 43 (24) |
| Baricitinib, no. (%) | 18 (10.1) |
| Corticosteroids, no. (%) | 65 (36.3) |
| Respiratory support, no. (%)<sup>†</sup> |  |
| Room air, no. (%) | 89 (49.7) |
| O2 nasal cannula, no. (%) | 20 (11.2) |
| O2 Venturi mask, no. (%) | 38 (21.2) |
| HFNC/CPAP/NIV, no. (%) | 8 (4.5) |
| MV, no. (%) | 23 (12.8) |
| Median length of MV, days [1<sup>st</sup>, 3<sup>rd</sup> quartile] | 13 [10.5, 25.5] |
| ECMO, no. (%) | 1 (0.6) |
| Outcomes and complications<sup>‡</sup> |  |
| Median length of hospital stay, days [1<sup>st</sup>, 3<sup>rd</sup> quartile] | 12 [9, 18] |
| ICU admission, no. (%)<sup>§</sup> | 34 (19) |
| Median length of ICU stay, days [1<sup>st</sup>, 3<sup>rd</sup> quartile] | 18.5 [11, 26] |
| Delirium, no. (%) | 8 (4.5) |
| Cerebrovascular event, no. (%) | 0 (0) |
| VTE, no. (%) | 17 (9.5) |
| Acute kidney injury, no. (%) | 9 (5) |
| Acute liver injury, no (%)<sup>¶</sup> | 59 (33) |
| Shock, no. (%) | 2 (1.1) |

Data are summarized as n (%) and medians [1<sup>st</sup>, 3<sup>rd</sup> quartile], as appropriate. ACE denotes angiotensin-converting enzyme, ECMO extracorporeal membrane oxygenation, HFNC/CPAP/NIV high-flow nasal cannula/continuous positive airway pressure/non-invasive ventilation, ICU intensive care unit, LDH lactate dehydrogenase, MV mechanical ventilation, SD standard deviation, SpO<sub>2</sub>/FiO<sub>2</sub> peripheral blood oxygen saturation/fraction of inspired oxygen, VTE venous thromboembolic event. \*Stage $\geq 2$ . <sup>†</sup>Maximum respiratory support needed during hospital stay. <sup>‡</sup>Complications were considered until the date of the interview administration. <sup>§</sup>Need for ICU admission at any time during hospitalization. <sup>||</sup> $\geq 2$ -fold increase of baseline serum creatinine or $\geq 50\%$ decrease in baseline glomerular filtration rate. <sup>¶</sup>Elevation of alanine transaminase (ALT) or aspartate transaminase (AST) enzymes $> 2 \times$ the upper limit of normal.

### Outpatient Clinic Assessment

Symptoms, radiological, and analytical findings were obtained. At the outpatient clinic visit, 58% of the patients had at least one or more persistent symptom, most frequently dyspnoea (22.9%). Up to 12.3% of patients reported memory complaints. More detailed descriptions of enduring symptoms are shown in eFigure 1.

41.9% of patients had an abnormal chest radiograph with sequelae or persistent infiltrates when seen at the outpatient clinic. Additional analytical parameters e.g. body mass index, and room air SpO2 are reported in eTable 2.

### Short-term Neurocognitive Function and Psychiatric Morbidity Assessment

For neurocognitive assessment, immediate verbal memory/learning, delayed verbal memory, semantic verbal fluency, and working memory were evaluated. From the full sample, 38% of patients presented moderate impairment and 11.2% severe impairment in immediate verbal memory. In relation to delayed memory, 11.8% of survivors had moderate impairment and 2.8% had severe impairment. In semantic verbal fluency, 34.6% of patients had moderate deficits and 8.4% severe deficits. Lastly, working memory was moderately impaired in 6.1% and severely impaired in 1.1% of survivors. The percentiles or scalar scores distribution for each neurocognitive function test are shown in eFigure 2. Finally, 105 (58.7%) patients met criteria for moderate neurocognitive impairment and 33 (18.4%) for severe neurocognitive impairment, as previously defined. Figure 2A details data regarding patients with affected neurocognitive tests.

**Figure 2.**
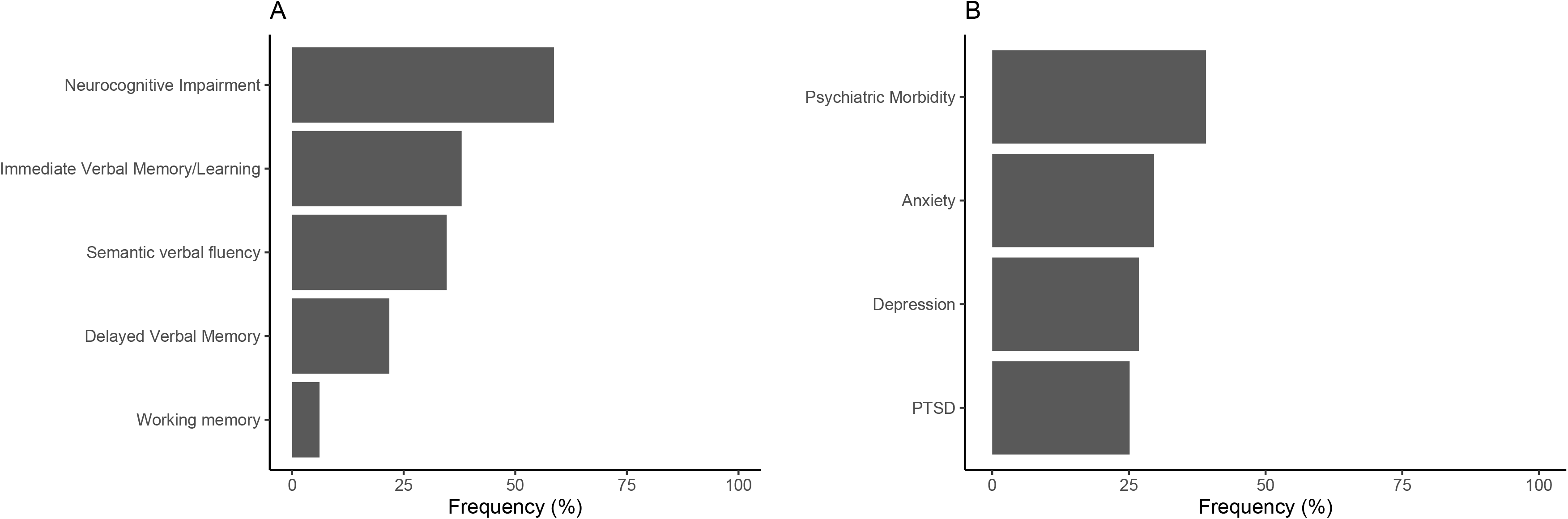
(A) Neurocognitive impairment and (B) psychiatric morbidity prevalence. PTSD denotes post-traumatic stress disorder.

The patients were also assessed for psychiatric morbidity, e.g. stress-related symptoms were analyzed. Of the 179 patients, a positive screening for anxiety was present in 29.6%, depression in 26.8%, and PTSD in 25.1% (Figure 2B). As previously defined, 39.1% of the patients screened positive for psychiatric morbidity. The scores distribution for the anxiety, depression, and PTSD questionnaires are shown in eFigure 3. Patients with stress-related symptoms presented a significantly higher prevalence of neurocognitive impairment compared with those without psychiatric symptoms (p<0.001) (eTable 3).

### Clinical Associations for Neurocognitive Impairment and Psychiatric Morbidity

The eTable 4 shows clinical data regarding neurocognitive impairment and psychiatric morbidity. Logistic regression models were performed to identify potential predictors for neurocognitive impairment and a positive screening for psychiatric morbidity (Table 2). A psychological trauma event prior to hospitalization due to COVID-19 was assessed as detailed in the Supplemental File. Trauma events were only identified in 17 patients. Of these, 3 had any PTSD symptoms in the past, but no patient reported long-lasting symptoms related to previous trauma at the time of assessment. For this reason, past history of traumatic event was not included in the models. Finally, delirium during the hospitalization and a positive screening for stress-related symptoms increased the odds of neurocognitive impairment approximately 4-fold. On the other hand, female gender and neurocognitive impairment diagnosis were related with a 2.5 and 4.56-fold increase respectively of the odds of psychiatric morbidity.

**Table 2.** Multivariable analysis for neurocognitive impairment and stress-related symptoms prediction.

| Neurocognitive impairment model |  |  |
| --- | --- | --- |
| <u>Variables</u> | <u>OR</u> | <u>95% Credibility Interval</u> |
| Any Comorbidity | 1.61 | 0.85-3.07 |
| SpO2/FiO2 | 1.01 | 1.0-1.01 |
| MV | 0.53 | 0.18-1.51 |
| **Delirium** | **4.05** | **1.03-16.4** |
| Acute kidney injury | 2.52 | 0.71-10.05 |
| **Positive screening for stress-related symptoms** | **3.97** | **2.06-7.80** |
| Stress-related symptoms model |  |  |
| <u>Variables</u> | <u>OR</u> | <u>95% Credibility Interval</u> |
| **Female gender** | **2.50** | **1.31-4.85** |
| **Neurocognitive impairment** | **4.56** | **2.27-9.47** |

MV denotes mechanical ventilation, OR odds ratio, SpO2/FiO2 peripheral blood oxygen saturation/fraction of inspired oxygen.

### Quality of Life and Functionality Assessment

The PCS and MCS scores for QoL appear on Figure 3A. The mean PCS was 42.5 (± 11.2) and the mean MCS was 45.5 (± 11.5). Briefly, 44.1% and a 39.1% of this cohort presented low QoL for PCS and MCS, respectively (Figure 3B). Only 1.1% and 9.5% presented high QoL for PCS and MCS, respectively. PCS and MCS scores did not significantly (p>0.05) correlate with each other (rho 0.030), and was also not correlated with age (rho −0.143 and rho 0.096) and years of education (rho 0.126 and rho 0.073). The eTable 5 describes the quality of life scores in the physical and mental components of neurocognitive and psychiatric domains. Patients with neurocognitive impairment or a positive screening for stress related symptoms showed lower scores for both PCS and MCS.

**Figure 3.**
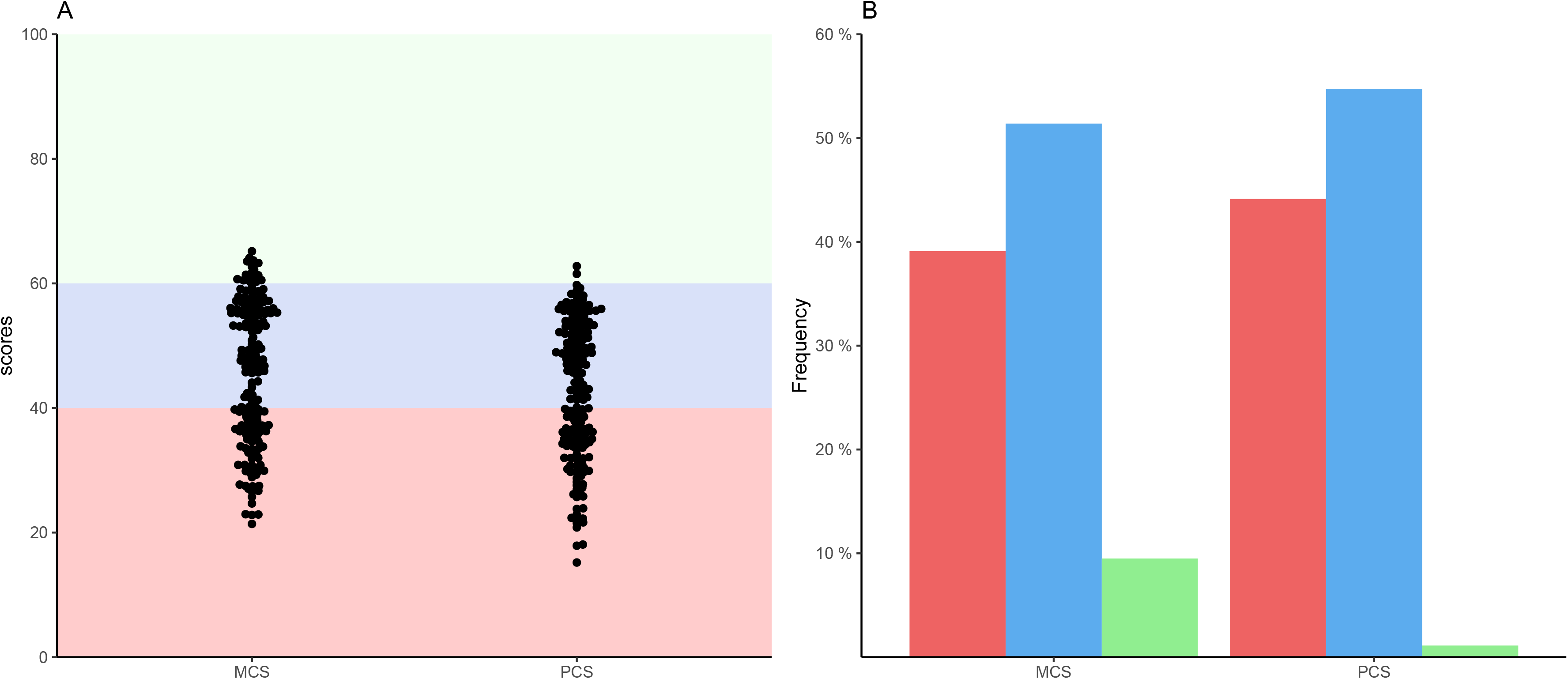
Quality of life. (A) Scores for Mental Component Summary (MCS) and Physical Component Summary (PSC). Green, blue, and red shaded areas denote high, normal, and low quality of life, respectively. (B) Quality of life distribution for MCS and PCS. The green, blue, and red bars denote high, normal, and low quality of life, respectively.

## DISCUSSION

To our knowledge, this is the first study to simultaneously evaluate the persistence of COVID-19 attributable symptoms, neurocognitive function, psychiatric symptoms, and QoL in mild to severe COVID-19 survivors requiring hospitalization after the acute phase. The main findings of the study are briefly summarized as follows: i) more than a half of survivors had long-lasting symptoms; ii) 58.7% presented at least moderate neurocognitive decline, 39.1% stress-related symptoms, and approximately 40% had poor QoL. The secondary findings were as follows: i) delirium during hospitalization and psychiatric comorbidity were associated with neurocognitive impairment; ii) and neurocognitive impairment and female gender were related to stress-related symptoms in the short-term.

Our study covers patients with different ranges of severity. Most of the available literature on neuropsychiatric consequences focused on critically ill patients with pneumonia, sepsis or ARDS who require intubation or admission to the ICU and there are no cohorts with similar severity to compare our results (15-20). The few studies in non-critically ill patients are focused on elderly patients. Moreover, previous studies have shown a declined neurocognitive function at hospital discharge in up to 100% of patients and up to 30% 1 year later (21-23). However, currently there is no data on neurocognitive function in the post-acute phase of COVID-19.

In the acute phase, several studies reported neurological problems like stroke, encephalitis, or delirium (6, 24). In a small study of COVID-19, one third of ICU survivors were found to have a dysexecutive syndrome (25). In this regard, follow-up studies in other coronaviruses outbreaks in the post-acute stage may be informative. In a recent review of 40 such studies after resolution of acute infection, Rogers et al reported substantial rates of memory impairment (44%) (5). Our findings show a high prevalence for neurocognitive impairment (58.7%), immediate verbal memory/learning (38%), delayed verbal memory (11.8%), semantic verbal fluency (34.6%), and executive function (6.1%) in a mild to severe cohort. Previous studies have found similar neurocognitive outcomes in critically-ill cohorts (17). In our study, delirium and having concurrent stress-related symptoms were linked to the odds of developing neurocognitive impairment. Delirium is a recognized risk factor for cognitive decline and reveals acute neurological damage that can persist over time (14, 15, 19). Furthermore, psychiatric disorders with coexisting stress-related symptoms such as PTSD and depression are associated with neurocognitive impairment (26). Screening for delirium and psychiatric morbidity may be useful to identify vulnerable subjects. No other convincing associations were found in the analysis, such as severity, levels of systemic inflammation, thrombosis related biomarkers, metabolic disorders, respiratory support, or oxygenation. There was also no association with age, although it should be noted that the neurocognitive tests were age-adjusted. Additional factors or mechanisms not explored in this study may further explain neurocognitive decline, such as neuroinvasion of SARS-CoV-2, endothelial injury, blood brain barrier disruption, or neuroinflammation (27).

In ICU survivors, similar rates of stress-related symptoms have been identified 3 and 12 months after discharge compared to our much less severe cohort (28, 29). In other coronavirus infections substantial rates of persistent depressed mood and anxiety have been reported (5). Recently, a study of COVID-19 survivors analyzed the presence of PTSD, depression, anxiety, obsessive-compulsive, and insomnia symptomatology (30). The authors reported very similar frequencies to those found in this study and showed that the female gender along with previous psychiatric diagnoses, but not systemic inflammation were associated with a higher prevalence of psychiatric symptoms. Indeed, women are two to three times more at risk to develop stress-related disorders (31). However, neurocognition and clinical data such as comorbidities, severity, or clinical outcomes (ICU admission, mechanical ventilation, delirium) were not characterized in that study.

Few studies about QoL in COVID-19 exist. In a recent report, worsened QoL was observed in 44.1% of patients discharged as measured with the EuroQoL visual analogue scale, defined as a 10-point different between the COVID-19 and post-acute stage visit (7). Our findings show that patients with neurocognitive impairment or psychiatric symptoms presented a poorer physical and mental quality of life. Both clinical problems have been associated with diminished quality of life in patients with psychiatric disorders and other diseases (32). Neurocognitive remediation and treatment of neuropsychiatric morbidity may improve QoL (33).

Our study has several limitations: 1) This is a single center study and the application of our findings to other populations should be extrapolated with caution, requiring external validation. 2) The tests were administered by telephone. The psychiatric screening for symptoms of anxiety, depression, and PTSD used self-reported measures after administering validated questionnaires. A face-to-face approach to the neurocognitive function and psychiatric morbidity may provide more precise information. 3) Although a drop-out rate of 21.8% is not likely to bias the present results, drop-outs might correspond to those patients with higher disabilities. If that were the case, the present findings would even underestimate the true prevalence of the outcomes. 4) Finally, the study lacked a control group to compare COVID-19 survivors with survivors of other acute diseases since most studies have focused on ICU survivors.

Study strengths not previously mentioned include the methodologically rigorous assessment of the outcomes. In previous studies, cognitive deficits were based on self-report (5). Therefore, no study has used neuropsychological tests to objectively establish cognitive deficits. All the selected tests were previously validated in the Spanish general population and included cut-off points for cognitive impairment or psychiatric screening. Moreover, confounding variables such as age and educational level were controlled in the case of cognitive performance. The study was performed in a well-characterized cohort and the high percentage of eligible patients who completed the assessment adds internal validation. Finally, very elderly patients and those with previous cognitive impairment or major psychiatric disorders were excluded. Further research on these more vulnerable patients are needed on the post-acute phase and in the long-term.

### Conclusions

In summary, our findings reveal a high prevalence of neurocognitive impairment, psychiatric comorbidity, and QoL in COVID-19 survivors after the acute phase, even in non-critically ill patients. These data have important clinical consequences and hence implications for health policies aimed to mitigate a wave of mental ill health following the current pandemic. Public health prevention, early detection, neurocognitive remediation, and treatment of psychiatric symptoms should be prioritized in COVID-19 survivors which may ultimately lead to an improvement in QoL and daily functioning.

## Data Availability

Data are accessible upon reasonable request

## DECLARATIONS

### Ethics Approval and Consent to Participate

This study was approved by the Ethics Committee of the Hospital Universitario y Politécnico La Fe (2020-280-1).

### Consent for Publication

All authors have accepted the publication of the manuscript.

### Availability of Data and Material

The datasets used and/or analyzed during the current study are available from the corresponding author on reasonable request.

### Authors’ Contributions

Conceptualization and study design: R. Méndez, V. Balanzá-Martínez, S.C. Luperdi, and R. Menéndez. Patient enrollment and database management: R. Méndez, P. González-Jiménez, L. Feced, L. Bouzas, K. Yépez, A. Ferrando, E. Zaldívar, and S. Reyes. Telephone interviews: I. Estrada. Statistical analysis: D. Hervás and R. Méndez. Drafting the manuscript: R. Méndez. Assistance in drafting the manuscript and critical review: V. Balanzá-Martínez, S.C. Luperdi, M. Berk, and R. Menéndez. Revision of manuscript and approval of the final version: all authors. R. Méndez and R. Menéndez are the guarantors.

## Acknowledgments

We are indebted to all patients and colleagues for their cooperation and assistance in this study. Always in our memory those who are no longer among us due to this pandemic.

